# Development and External Validation of a Mixed-Effects Deep Learning Model to Diagnose COVID-19 from CT Imaging

**DOI:** 10.1101/2022.01.28.22270005

**Authors:** Joshua Bridge, Yanda Meng, Wenyue Zhu, Thomas Fitzmaurice, Caroline McCann, Cliff Addison, Manhui Wang, Cristin Merritt, Stu Franks, Maria Mackey, Steve Messenger, Renrong Sun, Yitian Zhao, Yalin Zheng

## Abstract

**Objectives:** To develop and externally geographically validate a mixed-effects deep learning model to diagnose COVID-19 from computed tomography (CT) imaging following best practice guidelines and assess the strengths and weaknesses of deep learning COVID-19 diagnosis.

**Design:** Model development and external validation with retrospectively collected data from two countries.

**Setting:** Hospitals in Moscow, Russia, collected between March 1, 2020, and April 25, 2020. The China Consortium of Chest CT Image Investigation (CC-CCII) collected between January 25, 2020, and March 27, 2020.

**Participants:** 1,110 and 796 patients with either COVID-19 or healthy CT volumes from Moscow, Russia, and China, respectively.

**Main outcome measures:** We developed a deep learning model with a novel mixed-effects layer to model the relationship between slices in CT imaging. The model was trained on a dataset from hospitals in Moscow, Russia, and externally geographically validated on a dataset from a consortium of Chinese hospitals. Model performance was evaluated in discriminative performance using the area under the receiver operating characteristic (AUROC), sensitivity, specificity, positive predictive value (PPV), and negative predictive value (NPV). In addition, calibration performance was assessed using calibration curves, and clinical benefit was assessed using decision curve analysis. Finally, the model’s decisions were assessed visually using saliency maps.

**Results:** External validation on the large Chinese dataset showed excellent performance with an AUROC of 0.936 (95%CI: 0.910, 0.961). Using a probability threshold of 0.5, the sensitivity, specificity, NPV, and PPV were 0.753 (0.647, 0.840), 0.909 (0.869, 0.940), 0.711 (0.606, 0.802), and 0.925 (0.888, 0.953), respectively.

**Conclusions:** Deep learning can reduce stress on healthcare systems by automatically screening CT imaging for COVID-19. However, deep learning models must be robustly assessed using various performance measures and externally validated in each setting. In addition, best practice guidelines for developing and reporting predictive models are vital for the safe adoption of such models.

**Statements:** The authors do not own any of the patient data, and ethics approval was not needed. The lead author affirms that this manuscript is an honest, accurate, and transparent account of the study being reported, that no important aspects of the study have been omitted, and that any discrepancies from the study as planned (and, if relevant, registered) have been explained. Patients and the public were not involved in the study.

**Funding:** This study was funded by EPSRC studentship (No. 2110275), EPSRC Impact Acceleration Account (IAA) funding, and Amazon Web Services.

**Summary:** *What is already known on this topic:*

- Deep learning can diagnose diseases from imaging data automatically
- Many studies using deep learning are of poor quality and fail to follow current best practice guidelines for the development and reporting of predictive models
- Current methods do not adequately model the relationship between slices in CT volumetric data

*What this study adds:*

- A novel method to analyse volumetric imaging data composed of slices such as CT images using deep learning
- Model developed following current best-practice guidelines for the development and reporting of prediction models

## Introduction

Coronavirus disease 2019 (COVID-19) is an infectious respiratory disease caused by severe acute respiratory syndrome coronavirus 2 (SARS-CoV-2). Virus clinical presentation ranges from mild cold-like symptoms to severe viral pneumonia, which can be fatal.^1^ While some countries have achieved relative control through lockdowns, future outbreaks and new strains are expected to continue, with many experts believing the virus is here to stay.^2^ Detection and isolation is the most effective way to prevent further spread of the virus. Even with effective vaccines becoming widely available, with the threat of continued waves and new potentially vaccine-resistant variants, it is vital to further develop diagnostic tools for COVID-19. These tools will likely also apply to future outbreaks of other similar diseases as well as common diseases such as pneumonia.

The diagnosis of COVID-19 is usually determined by Reverse Transcription Polymerase Chain Reaction (RT-PCR), but this is far from being a gold standard. A negative test does not necessarily indicate a negative diagnosis, with one recent review finding that RT-PCR has a real-world sensitivity of around 70% and a specificity of 95%.^3^ Furthermore, an individual patient data systematic review^4^ found that RT-PCR often fails to detect COVID-19, and early sampling is key to reducing false negatives. Therefore, these tests are often more helpful to rule in COVID-19 rather than ruling out. If a patient presents with symptoms of COVID-19, but an RT-PCR test is negative, then further tests are often required.^1^ Consecutive negative tests with at least a one-day gap are recommended; however, this still does not guarantee that the patient is negative for COVID-19.^5^ Computed tomography (CT) can play a significant role in diagnosing COVID-19.^6^ Given the excessive number of COVID-19 cases worldwide and the strain on resources expected, automated diagnosis might reduce the burden on reporting radiologists.

CT images are made up of many slices, creating a three dimensional (3D)-like structure. Previous approaches, such as those used by Li et al.^7^ and Bai et al.,^8^ treat the image as separate slices and use a pooling layer to concatenate the slices. An alternative approach assumes the slices form a 3D structure and use a 3D CNN, such as that proposed in CoviNet.^9^ A fundamental limitation of these methods is the need for the same number of slices as their inputs, but the number of slices often varies between different CT volumes. Instead, we propose using a novel mixed-effects layer to consider the relationship between slices in each scan. Mixed-effects models are commonly used in traditional statistics,^10 11^ but we believe this is the first time that mixed-effects models have been utilised in such a way. It has been observed that some lobes of the lung are more often affected by COVID-19 than others^12 13^ with lower lobe distribution being a prominent feature of COVID-19,^14^ the fixed-effects take this into account by considering where each slice is located within the scan.

Deep learning has shown great potential in the automatic classification of disease, often achieving expert-level performance. Such models could screen and monitor COVID-19 by automatically analysing routinely collected CT images. As observed by Wynants et al.^15^ and Roberts et al.,^16^ many models are already developed to diagnose COVID-19, which often obtain excellent discriminative performance; however, very few of these models, if any, are suitable for clinical use, mainly due to a lack of robust analysis and reporting. These models often suffer from common pitfalls, making them unsuitable for broader adoption. Roberts et al.^16^ identified three common areas in which models often fail these are: (1) a lack of adequately documented methods for reproducibility, (2) failure to follow established guidelines and best practices for the development of deep learning models, and an absence of external validation displaying the model’s applicability to a broader range of data outside of the study sample. Failure to address these pitfalls leads to profoundly flawed and biased models, making them unsuitable for deployment.

In this work, we aim to address the problems associated with previous models by following guidelines for the reporting^17 18^ and development^19^ of prediction models to ensure that we have rigorous documentation allowing the methods developed here to be replicated. In addition, we will make code and the trained model publicly available at [github.com/JTBridge/ME-COVID19] to promote reproducible research and facilitate adoption. Finally, we use a second dataset from a country other than the development dataset to externally validate the model and report a range of performance measures evaluating the model’s discrimination, calibration, and clinical usefulness.

Hence, our main aim is to develop a mixed-effects deep learning model to accurately classify images as healthy or COVID-19, following best practice guidelines. Our secondary aim is to show how deep learning predictive algorithms can satisfy current best practice guidelines to create reproducible and less biased models.

## Methods

Our proposed method consists of a feature extractor and a two-stage generalised linear mixed-effects model (GLMM),^20^ with all parameters estimated within the deep learning framework using backpropagation. First, a series of CT slices forming a CT volume is input to the model. In our work, we use 20 slices. Next, a convolutional neural network (CNN) extract relevant features from the model and creates a feature vector for each CT slice. Then, a mixed-effects layer concatenates the feature vectors into a single vector. Finally, a fully connected layer followed by a sigmoid activation gives a probability of COVID-19 for the whole volume. The mixed effects and fully connected layer with sigmoid activation are analogous to a linear GLMM in traditional statistics. The overall framework is shown in Figure 1.

**Figure 1:**
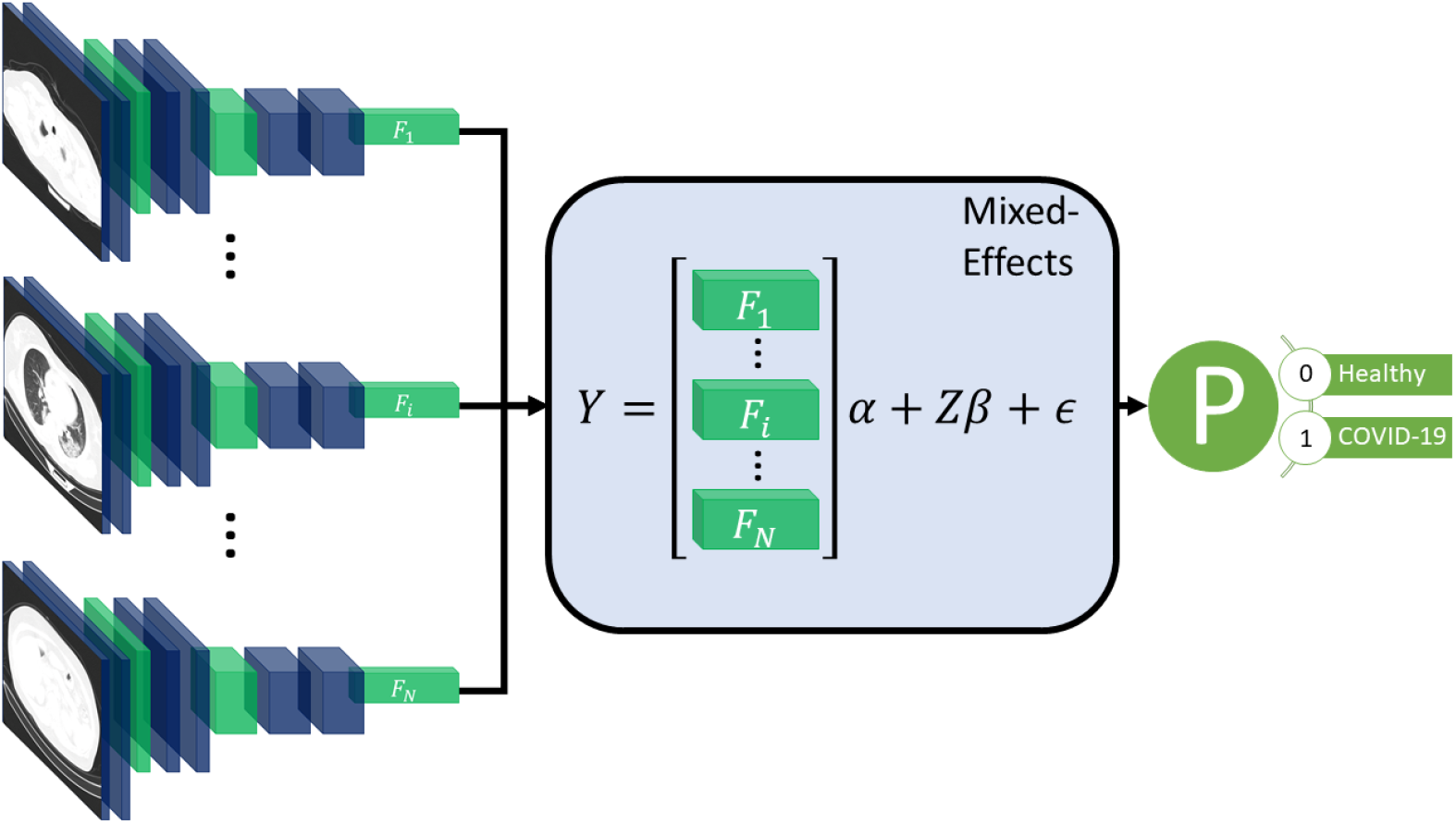
**Diagram of t**he overall framework. Twenty slices are chosen from a CT volume. Each slice is fed into a CNN with shared weights, which outputs a feature vector of length 2048 for each image. The feature vectors form a 20-by-2048 fixed effects matrix, X, for the GMM model with a random-effects matrix, Z, consisting of an identity matrix. A mixed-effects model is used to model the relationship between slices. Finally, a fully connected layer and sigmoid activation return a probability of the diagnosis.

**Figure 2:**
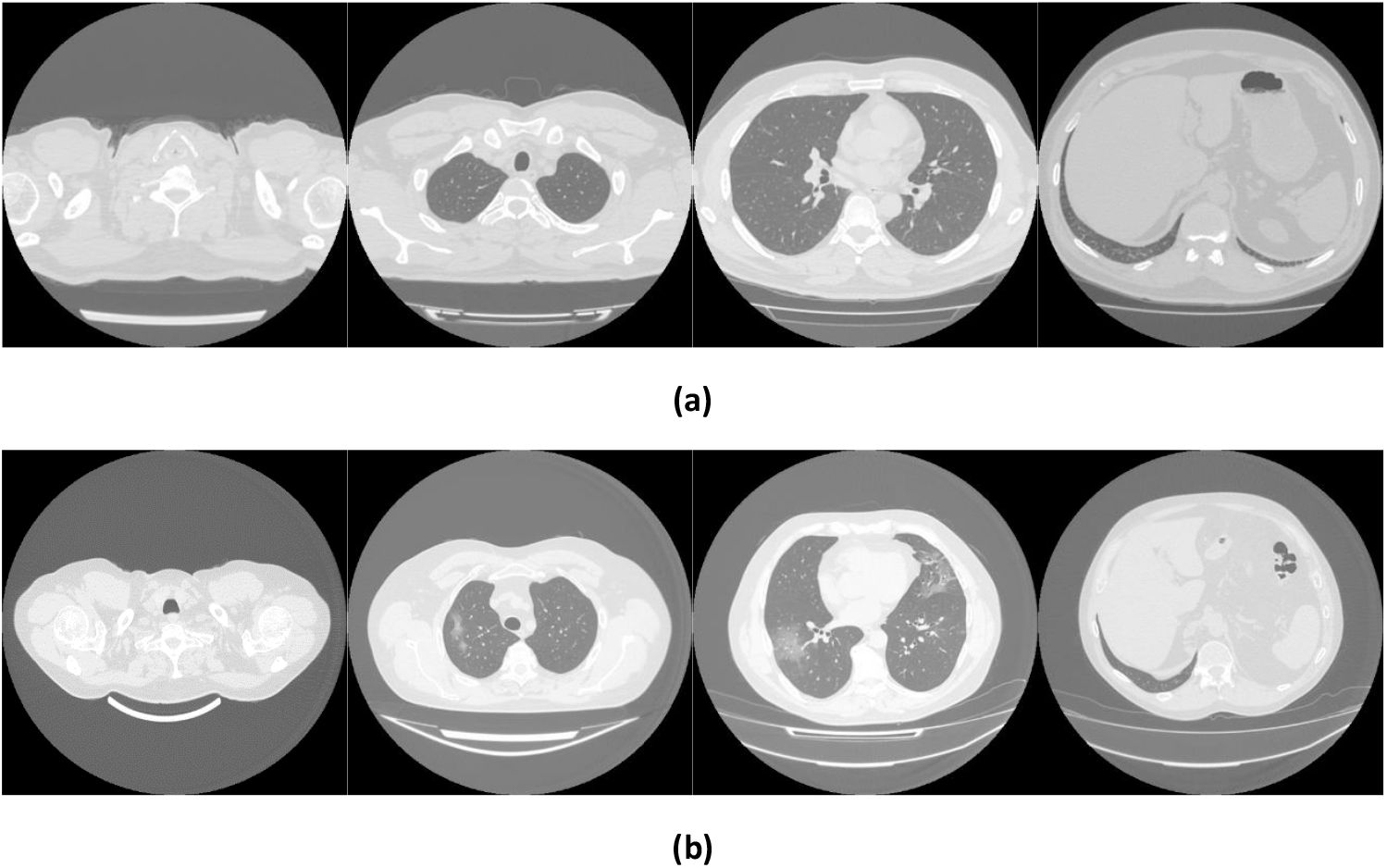
Example images showing (a) healthy and (b) COVID-19 lungs taken from the Mosmed dataset.

### Feature extractor

For the feature extractor, we use a CNN. In this work, we chose InceptionV3^21^ as it is relatively efficient and commonly used. InceptionV3 outputs a feature vector of length 2048. To reduce the time needed to reach convergence, we pretrained the CNN on ImageNet.^22^ A CNN is used for each slice, with shared weights between CNNs; this reduces the amount of computational power required. Following the CNN, we used a global average pooling layer to reduce each image to a feature vector for each slice. We then added a dropout of 0.6 to improve generalizability to unseen images. We form the feature vectors into a matrix of shape 20 × 2048. Although we used InceptionV3^21^ here, other networks would also work and may provide better performance on other similar tasks. We then need to concatenate these feature vectors into a single feature vector for the whole volume; normally, pooling is used, in our work we propose using a mixed-effects models.

### Mixed-effects network

We propose utilising a novel mixed-effects layer to model the relationship between slices. Mixed-effects models are a statistical model consisting of a fixed-effects part and a random-effects part. The fixed-effects part models the relationship within the CT slice; the random effects can model the spatial correlation between CT slices within the same image.^11^ For volumetric data, the number of slices may differ significantly due to various factors such as imaging protocol and the size of the patient. Some volumes may have fewer images than the model is designed to use, which leads to missing data. Mixed-effects models can deal with missing data provided the data are missing at random. The mixed-effects model is given by

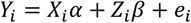

where *Y*_*i*_, *X*_*i*_, *Z*_*i*_, *e*_*i*_ are vectors of outcomes, fixed effects design matrix of shape 20 × 2048, random effects design matrix of shape 20 × 20, and vector of error unknown random errors of the *i*th patient of shape 20, respectively, and *α, β* are fixed and random effects parameters, both of length 20. We assume that the random effects *β* are normally distributed with 0 and variance *G*

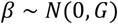

We also assume independence between the random effects and the error term.

The fixed effects design matrix, *X*, is made up of the feature vectors output from the feature extraction network. For the random effects design matrix, *Z*, we use an identity matrix with the same size as the number of slices; in our experiments, this is 20. The design matrix is then given by

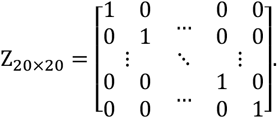

This matrix easily generalises to any number of slices. If the distance between slices is not uniform, the values can be altered accordingly. We assumed no particular correlation matrix. We included the fixed and random intercept in the model. All parameters for the mixed-effects layer were initialised using the Gaussian distribution with mean 0 and standard deviation 0.05.

A type of mixed-effects modelling has previously been combined with deep learning for gaze estimation.^23^ However, their mixed-effects method is very different from our proposed method; they used the same design matrix for fixed and random effects. In addition, they also estimated random-effects parameters with an expectation-maximisation algorithm, which was separate from the fixed effects estimation, which used deep learning. In our work, we utilise a spatial design matrix to model the spatial relationship between slices and estimate parameters within the deep learning framework using backpropagation without the need for multiple stages.

### Loss function

As the parameters in the model are all estimated using backpropagation, we must ensure that the assumption of normally distributed random effects parameters with mean zero is valid. We achieve this by introducing a loss function for the random effects parameters, which enforces a mean, skewness, and excess kurtosis of 0. We measure skewness using the adjusted Fisher–Pearson standardised moment coefficient

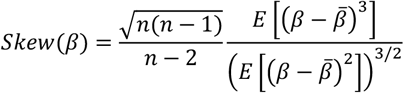

and the excess kurtosis using

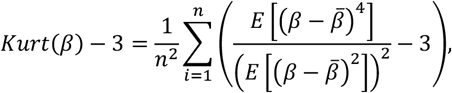

where *n* is the length of *β*, 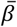 is the mean of *β* and *E*[.] is the expectation function. The Gaussian distribution has a kurtosis of 3; therefore, the excess kurtosis is given by the kurtosis minus 3. This formula for this fixed-effects parameters loss function which we aim to minimise, is then given by

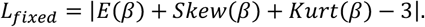

For the classification, we use the Brier Score^24^ as the loss function, which is given by

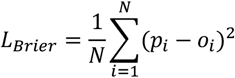

where *N* is the total number of samples, *p*_*i*_ is the predicted probability of sample *i* and *o*_*i*_ is the observed outcome of sample *i*. The Brier score is the same as the mean squared error of the predicted probability.

We chose to use the Brier Score over the more commonly used binary cross-entropy because it can be decomposed into two components: refinement and calibration. Calibration is often overlooked in deep learning models but is vital to assess the safety of any prediction model. The refinement component combines the model’s resolution and uncertainty and measures the model’s discrimination. The calibration component can be used as a measure of the model calibration. Therefore, the Brier Score can be used to optimize both the discrimination and calibration of the model. The overall loss function is given by

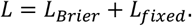

A scaling factor could be introduced to weight one part of the loss function as more important than the other; however, we give both parts of the loss function equal weighting in our work.

We also transformed the labels as suggested by Platt^25^ to reduce overfitting. The negative and positive labels become

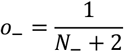

And

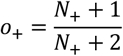

respectively, where *N*_−_ and *N*_+_ are the total number of negative and positive cases in the training set. This is similar to label smoothing as commonly used in deep learning, but the new targets are chosen by applying Baye’s Rule to the out-of-sample data to prevent overfitting.

### Classification layer

The output of the mixed-effects layer is a single vector, which is the same length as the number of slices used. For example, in our work, we had a vector of length 20. Furthermore, we used a fully connected layer with sigmoid activation to obtain a probability of the scan showing COVID-19; the sigmoid activation is analogous to the logistic link function in traditional statistics. Finally, we added an L1 regularisation term of 0.1 and an L2 regularisation term of 0.01 to the kernel to reduce overfitting.

### Model performance

Many deep learning models focus on assessing discriminative performance only, using measures such as the area under the receiver operating characteristic curve (AUROC), sensitivity, and specificity. To better understand the model performance and impact, we report performance measures in three broad areas: discrimination, calibration, and clinical usefulness.^26^ Discrimination assesses how well a model can discriminate between healthy and COVID-19 positive patients. Models with excellent discriminative performance can still produce unreliable results, with vastly overestimated probabilities regardless of the true diagnosis.^27^ Model calibration is often overlooked and rarely reported in deep learning, if at all; however, poorly calibrated models can be misleading and lead to dangerous clinical decisions.^27^ Calibration can be assessed using four levels, with each level indicating better calibration than the last.^28^ The fourth and most stringent level (strong calibration) requires the correct model to be known, which in turn requires predictors to be non-continuous, and an infinite amount of data to be used and is therefore considered utopic. We consider the third level (moderate calibration) using calibration curves. Moderate calibration will ensure that the model is at least not clinically harmful. Finally, measures of clinical usefulness assess the clinical consequences of the decision and acknowledge that a false positive may be more or less severe than a false negative.

Firstly, the discriminative performance is assessed using AUROC using the pROC package in R,^29^ with confidence intervals constructed using DeLong’s^30^ method. For sensitivity, specificity, positive predictive value (PPV), and negative predictive value (NPV), we use the epiR^31^package in R;^29^ with 95% confidence intervals constructed using Jeffrey’s prior.^32^ We report performance at a range of probability thresholds to demonstrate how the thresholds can be adjusted to reduce false positives or false negatives depending on the setting.^33^ Secondly, we assess model calibration using calibration curves created using the CalibrationCurves package,^28^ which is based on the rms^34^ package. Finally, we assess the clinical usefulness of the model using decision curve analysis.^35^ Net benefits are given at various thresholds, and models which reach zero net benefit at higher thresholds are considered more clinically useful. Two brief sensitivity analyses are performed, one assessing the model’s ability to deal with missing data and the other assessing its ability to deal with noise. To improve the model’s interpretability and reduce the black-box nature, we produce saliency maps^36^ that show which areas of the image are helpful to the model in the prediction. We also check the assumption of normally distributed random-effects parameters.

### Comparison models

To assess the added benefit of using our mixed-effects method, we compare against networks that use alternative methods. Both COVNet^7^ and a method proposed by Bai et al.^8^ propose deep learning models that consider the slices separately before concatenating the features using max pooling. COVNet uses a ResNet50^37^ CNN to extract features and pooling layers to concatenate the features before a fully connected classification layer. The model proposed by Bai et al. uses EfficientNetB4^38^ to extract features followed by a series of full-connected layers with batch normalisation and dropout; average pooling is then used to concatenate the feature vectors before classification. While max pooling is simple and computationally efficient, it cannot deal with pose variance and does not model the relationship between slices.

An alternative method to pooling is treating the scans as 3D, such as in CoviNet^39^. CoviNet takes the whole scan and uses a 16 layer 3D CNN followed by pooling and fully connected layers. We implemented these models as described in their respective papers.

In all comparison experiments, we kept hyperparameters, such as learning rate, learning rate decay, and data augmentation, the same to ensure the comparisons were fair. For COVNet^7^ and the model proposed by Bai et al.,^8^ we pretrained the CNNs on ImageNet as they also did; however, no pretrained models were available for CoviNet. For the loss function, we also used the Brier score.^24^

### Computing

Models were developed using an Amazon Web Services p3.8xlarge node with four Tesla V100 16GiB GPUs and 244GiB available memory. Model inference was performed on a local Linux machine running Ubuntu 18.04, with a Titan X 12GiB GPU and 32GiB available memory. Model development and inference were performed using Tensorflow 2.4,^40 41^ and R 4.0.5 ^29^ was used to produce evaluation metrics^42 43^ and graphs.^34 44^ We used mixed precision to reduce the computational cost, which uses 16-bit floating-point precision in all layers, except for the mixed-effects and classification layers, where 32-bit floating-point precision is used.

We used the Adam optimiser^45^ with an initial learning rate of 1e-4; if the internal validation loss did not improve for three epochs, we reduced the learning rate to 20%. In addition, we assumed convergence and stopped training if the loss did not improve for ten epochs to reduce the time spent training and the energy used.

### Data

There is currently no established method for estimating the sample size estimate in deep learning. We propose treating the final fully connected classification layer as the model and treating previous layers as feature extraction. We can then use the number of parameters in the final layer to estimate the required sample size. Using the ‘pmsampsize’ package^46^ in R, we estimate the required minimum sample size in the development set. We use a conservative expected C-statistic of 0.8, with 21 parameters and an estimated disease prevalence of 80% based on datasets used in other studies. This gives a minimum required sample size of 923 patients in the training set. For model validation, around 200 patients with the disease and 200 patients without the disease are estimated to be needed to assess calibration.^28^

All data used here is retrospectively collected and contains hospital patients with CT scans performed during the COVID-19 pandemic. The diagnosis was determined by examining radiological features of the CT scan for signs of COVID-19, such as ground-glass opacities. For model development, we use the MosMed dataset,^47^ which consists of a total of 1,110 CT scans displaying either healthy or COVID-19 infected lungs. The scans were performed in Moscow hospitals between March 1, 2020, and April 25, 2020. We split the dataset into two sets for training and internal validation on the patient level. The training set is used to train the model, and the internal validation set is used to select the best model based on the loss at each epoch; this helps prevent overfitting on the training set. In addition, we obtained images from a publicly available dataset published by Zhang et al.^48^ consisting of CT images from a consortium of Chinese hospitals.

Overall, this allows us to perform external geographical validation in another country and to better evaluate the developed model. In addition, we will be able to assess how well a deep learning model generalises to other populations. A summary of all the datasets used is shown in Table 1. We have 923 patients in the training set and at least 200 patients in each class for the external validation set.

**Table 1:** Summary of the datasets used.

| Dataset | Location | Use | Healthy/COVID19 |
| --- | --- | --- | --- |
| MosMed Training | Moscow, Russia | Training | 169/856 |
| MosMed Validation | Moscow, Russia | Internal Validation | 85/285 |
| Zhang et al.<sup>48</sup> | China | External Validation | 243/553 |

### Data pre-processing and augmentation

The MosMed dataset was converted from Dicom image format into PNG, normalised to have a mean of 120 and a variance of 95. Images were ordered from the top of the lungs to the bottom. During training, we applied random online data augmentation to the images. This alters the image slightly and gives the effect of increasing the training dataset size, although this is not as good as expanding the training dataset with more samples. First, we adjusted the brightness and contrast between 80% and 120%. We then rotated the image plus or minus 5 degrees and cropped the image up to 20% on each side. Finally, we flipped the image horizontally and vertically with a probability of 50% each. All random values were chosen using the uniform distribution except for the flips, which were chosen using a random bit.

The dataset taken from Zhang et al.^48^ required a large amount of sorting to be made suitable for use. Some of the scans were pre-segmented and only showed the lung areas, while others showed the whole CT scan. We removed any pre-segmented images. Identifying information on some images had to be cropped to reduce bias in the algorithm. In addition, many of the scans were duplicates but were not labelled as such, and many scans were incomplete, only showing a few lung slices or not showing any lung tissue at all. We only used complete scans with one scan per patient. Finally, some scans needed to be ordered top to bottom. Using the bilinear sampling algorithm, all images were resized to 256 by 256 pixels, and image values were divided by 255 to normalise between 0 and 1.

The MosMed dataset has a median of 41 slices, a minimum of 31 slices and a maximum of 72 slices. The Zhang et al. dataset has much greater variability in scan size with a median of 61 slices, a minimum of 19 slices, and a maximum of 415 slices. We present histograms showing the number of slices per scan in Figure 3. We require a fixed number of slices as input, and we chose 20 as the slice size. For all scans, we included the first and last images. If scans had more than 20 slices, we sampled uniformly to select 20. Only one scan in the Zhang et al. dataset had less than 20 slices; a blank slice replaced this slice; the mixed-effects model can account for missing data.

**Figure 3:**
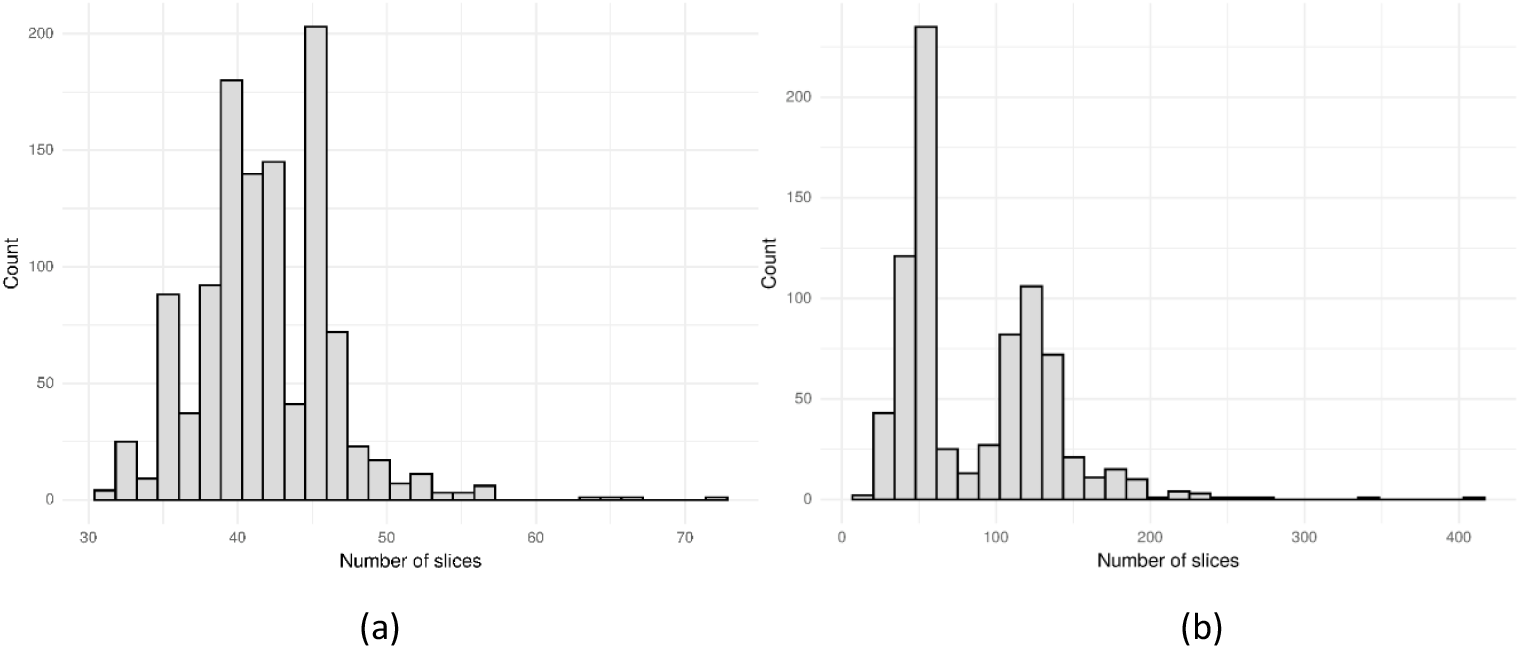
Histogram showing the number of slices per scan for (a) the MosMed^47^ dataset and (b) the Zhang et al.^48^ dataset. The MosMed dataset has much fewer slices on average with a much smaller spread.

**Figure 4:**
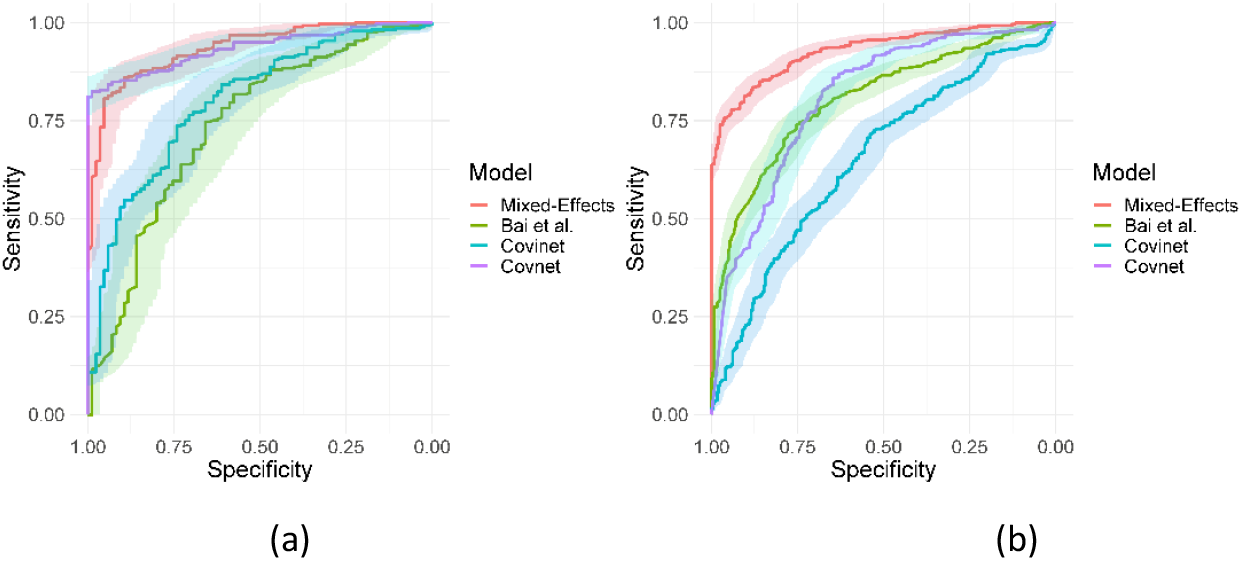
Receiver operating characteristic curves for (a) the MosMed internal validation set and (b) the Zhang et al.^48^ external validation set.

## Results

On the internal validation dataset, the proposed model attained an AUROC of 0.936 (95%CI: 0.910, 0.961). Using a probability threshold of 0.5, the sensitivity, specificity, NPV, and PPV were 0.753 (0.647, 0.840), 0.909 (0.869, 0.940), 0.711 (0.606, 0.802), and 0.925 (0.888, 0.953), respectively. The model proposed by Bai et al.^8^ attained an AUROC of 0.731 (0.674, 0.80). However, despite attaining a reasonably AUC value, the model was badly calibrated, and the predicted probabilities of COVID-19 were all clustered around 0.42, meaning that the sensitivity, specificity, PPV, and NPV are meaningless. We tried to retrain the model and rechecked the code implementation; however, we could not obtain more meaningful results. Covinet^9^ attained an AUROC of 0.810 (0.748, 0.853). Using a probability threshold of 0.5, the sensitivity, specificity, NPV, and PPV were 0.824 (0.726, 0.898), 0.596 (0.537, 0.654), 0.378 (0.308, 0.452), and 0.919 (0.870 0.954), respectively. COVNet^7^ attained an AUROC of 0.935 (0.912, 0.959). Using a probability threshold of 0.5, the sensitivity, specificity, NPV, and PPV were 1.0 (0.958, 1.0), 0.796 (0.745, 0.842), 0.594 (0.509, 0.676), and 1.0 (0.984, 1.0), respectively. Full results for a range of probability thresholds are shown in Table 2.

**Table 2:** Area under the receiver operating characteristic curve (AUROC), sensitivity, specificity, positive predictive value (PPV) and negative predictive value (NPV) on the internal validation dataset. Point estimates and 95% confidence intervals were calculated using De Long’s method for AUROC and Jeffrey’s interval for sensitivity, specificity, PPV, and NPV. Results are shown at a range of probability thresholds.

| Model | AUROC | Threshold | Sensitivity | Specificity | PPV | NPV |
| --- | --- | --- | --- | --- | --- | --- |
| Bai et al | 0.731<br>(0.674, 0.80) | 0.3 | 0.0 (0.0, 0.042) | 1.0 (0.987, 1.0) | NA | 0.77 (0.724, 0.812) |
|  |  | 0.4 | 0.012 (0, 0.064) | 0.996 (0.981, 1.0) | 0.50 (0.013, 0.987) | 0.772 (0.725, 0.814) |
|  |  | 0.5 | 1.0 (0.958, 1.0) | 0.0 (0.0, 0.013) | 0.230 (0.188, 0.276) | NA |
|  |  | 0.6 | 1.0 (0.958, 1.0) | 0.0 (0.0, 0.013) | 0.230 (0.188, 0.276) | NA |
|  |  | 0.7 | 1.0 (0.958, 1.0) | 0.0 (0.0, 0.013) | 0.230 (0.188, 0.276) | NA |
| CoviNet | 0.801<br>(0.748, 0.853) | 0.3 | 0.459 (0.350, 0.570) | 0.898 (0.857, 0.931) | 0.574 (0.448, 0.693) | 0.848 (0.802, 0.886) |
|  |  | 0.4 | 0.706 (0.597, 0.80) | 0.761 (0.708, 0.810) | 0.469 (0.380, 0.559) | 0.897 (0.851, 0.932) |
|  |  | 0.5 | 0.824 (0.726, 0.898) | 0.596 (0.537, 0.654) | 0.378 (0.308, 0.452) | 0.919 (0.870, 0.954) |
|  |  | 0.6 | 0.918 (0.838, 0.966) | 0.446 (0.387, 0.505) | 0.331 (0.271, 0.394) | 0.948 (0.895, 0.979) |
|  |  | 0.7 | 0.965 (0.90, 0.993) | 0.246 (0.197, 0.30) | 0.276 (0.226, 0.331) | 0.959 (0.885, 0.991) |
| CovNet | 0.935<br>(0.912, 0.959) | 0.3 | 0.941 (0.868, 0.981) | 0.839 (0.791, 0.879) | 0.635 (0.544, 0.719) | 0.98 (0.953, 0.993) |
|  |  | 0.4 | 0.965 (0.90, 0.993) | 0.825 (0.775, 0.867) | 0.621 (0.533, 0.704) | 0.987 (0.964, 0.997) |
|  |  | 0.5 | 1.0 (0.958, 1.0) | 0.796 (0.745, 0.842) | 0.594 (0.509, 0.676) | 1.0 (0.984, 1.0) |
|  |  | 0.6 | 1.0 (0.958, 1.0) | 0.779 (0.726, 0.826) | 0.574 (0.490, 0.655) | 1.0 (0.984, 1.0) |
|  |  | 0.7 | 1.0 (0.958, 1.0) | 0.761 (0.708, 0.810) | 0.556 (0.473, 0.636) | 1.0 (0.984, 1.0) |
| Mixed-Effects<br>(Ours) | 0.936<br>(0.910, 0.961) | 0.3 | 0.588 (0.476, 0.694) | 0.961 (0.932, 0.981) | 0.820 (0.70, 0.906) | 0.887 (0.846, 0.920) |
|  |  | 0.4 | 0.659 (0.548, 0.758) | 0.933 (0.898, 0.959) | 0.747 (0.633, 0.840) | 0.902 (0.862, 0.933) |
|  |  | 0.5 | 0.753 (0.647, 0.840) | 0.909 (0.869, 0.940) | 0.711 (0.606, 0.802) | 0.925 (0.888, 0.953) |
|  |  | 0.6 | 0.812 (0.712, 0.888) | 0.884 (0.841, 0.919) | 0.676 (0.577, 0.766) | 0.940 (0.905, 0.960) |
|  |  | 0.7 | 0.906 (0.823, 0.958) | 0.832 (0.783, 0.873) | 0.616 (0.525, 0.702) | 0.967 (0.937, 0.986) |

Calibration curves in Figure 5 show reasonable calibration for the mixed-effects model, although the model may still benefit from some recalibration. The other models do not have good calibration and likely provide harmful predictions. The decision curve in Figure 6 shows that the proposed model is of great clinical benefit compared to the treat all and treat-none approach.

**Figure 5:**
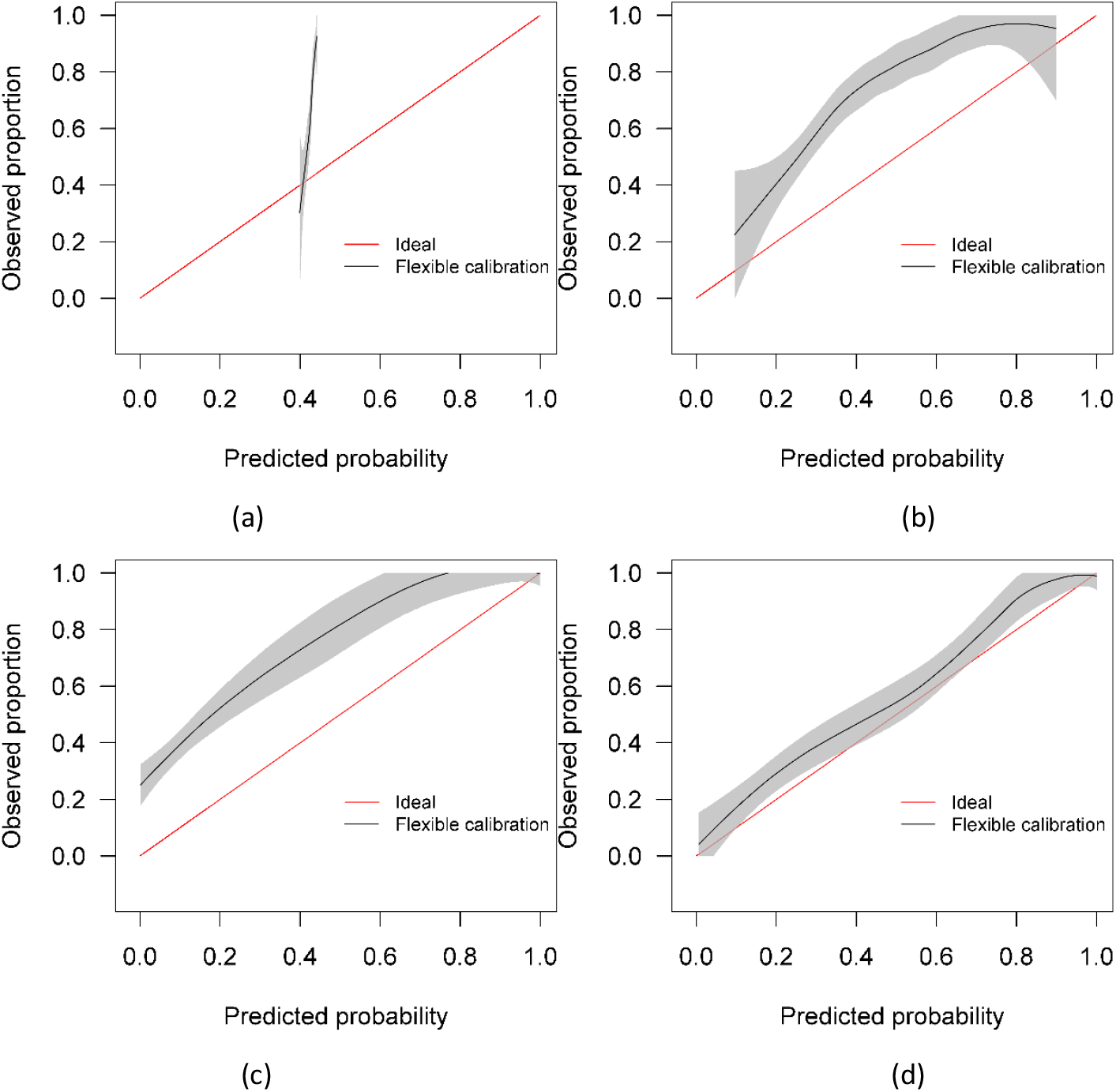
Calibration curves for (a) the Bai et al.^8^ model (b) the Covinet model^9^, (c) the Covnet model^7^, (d) the proposed mixed-effects model on the Mosmed internal validation dataset.

**Figure 6:**
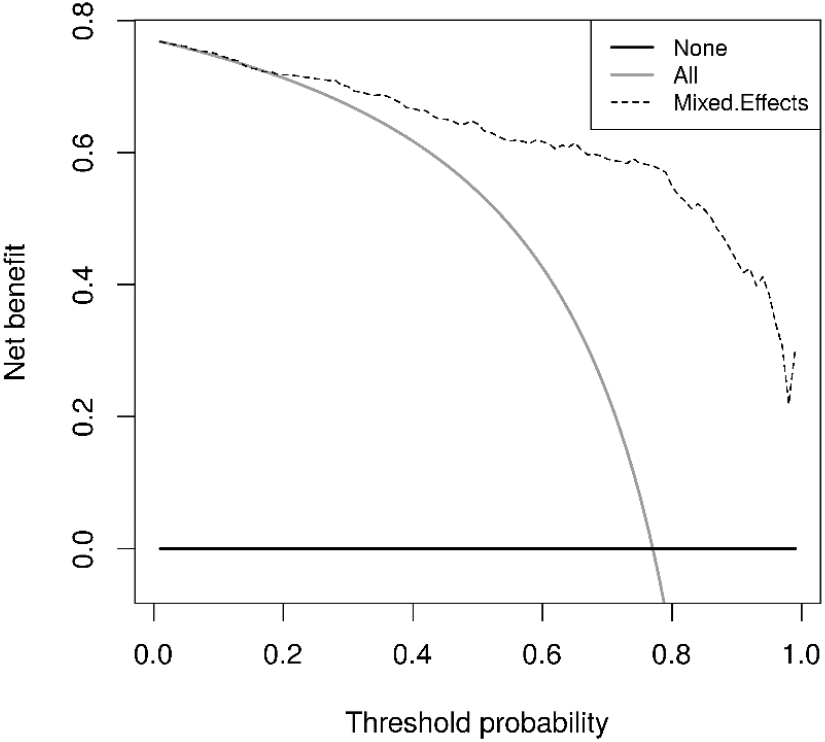
Decision curves for the proposed mixed-effects model on the Mosmed internal validation dataset.

It is important to remember that the model was selected using this internal testing set to avoid overfitting on the training set; therefore, these results are biased, and the external validation results are more representative of the true model performance.

On the external geographical validation dataset, the proposed model attained an AUROC of 0.930 (0.914, 0.947). With a probability threshold of 0.5, the sensitivity, specificity, NPV, and PPV were 0.778 (0.720, 0.828), 0.882 (0.853, 0.908), 0.744 (0.686, 0.797), and 0.90 (0.872, 0.924), respectively. The model proposed by Bai et al.^8^ again attained a reasonable AUROC of 0.805 (0.774, 0.836); however, the sensitivity, specificity, NPV, and PPV were meaningless. Covinet^9^ attained an AUROC of 0.651 (0.610, 0.691). Using a probability threshold of 0.5, the sensitivity, specificity, NPV, and PPV were0.008 (0.001, 0.029), 0.991 (0.979, 0.997), 0.286 (0.037, 0.710), and 0.695 (0.661, 0.727), respectively. COVNet^7^ attained an AUROC of 0.808 (0.775, 0.841). With a cut-off point of 0.5, the sensitivity, specificity, NPV, and PPV were 0.387 (0.325, 0.451), 0.940 (0.917, 0.959), 0.740 (0.655, 0.814), and 0.777 (0.744, 0.808), respectively. Full results are shown in Table 3.

**Table 3:** Area under the receiver operating characteristic curve (AUROC), sensitivity, specificity, positive predictive value (PPV) and negative predictive value (NPV) on the external validation dataset. Point estimates and 95% confidence intervals were calculated using De Long’s method for AUROC and Jeffrey’s interval for sensitivity, specificity, PPV, and NPV. Results are shown at a range of probability thresholds.

| Model | AUROC | Threshold | Sensitivity | Specificity | PPV | NPV |
| --- | --- | --- | --- | --- | --- | --- |
| Bai et al | 0.805<br>(0.774, 0.836) | 0.3 | 0.0 (0.0, 0.015) | 1.0 (0.993, 1.0) | NA | 0.695 (0.661, 0.727) |
|  |  | 0.4 | 0.0 (0.0, 0.015) | 1.0 (0.993, 1.0) | NA | 0.695 (0.661, 0.727) |
|  |  | 0.5 | 1.0 (0.985, 1.0) | 1.0 (0.0, 0.007) | 0.305 (0.273, 0.339) | NA |
|  |  | 0.6 | 1.0 (0.985, 1.0) | 1.0 (0.0, 0.007) | 0.305 (0.273, 0.339) | NA |
|  |  | 0.7 | 1.0 (0.985, 1.0) | 1.0 (0.0, 0.007) | 0.305 (0.273, 0.339) | NA |
| CoviNet | 0.651<br>(0.610, 0.691) | 0.3 | 0.0 (0.0, 0.015) | 1.0 (0.993, 1.0) | NA | 0.695 (0.661, 0.727) |
|  |  | 0.4 | 0.0 (0.0, 0.015) | 1.0 (0.993, 1.0) | NA | 0.695 (0.661, 0.727) |
|  |  | 0.5 | 0.008 (0.001, 0.029) | 0.991 (0.979, 0.997) | 0.286 (0.037, 0.710) | 0.695 (0.661, 0.727) |
|  |  | 0.6 | 0.160 (0.117, 0.213) | 0.929 (0.905, 0.949) | 0.50 (0.385, 0.615) | 0.716 (0.681, 0.749) |
|  |  | 0.7 | 0.551 (0.487, 0.615) | 0.694 (0.654, 0.733) | 0.442 (0.385, 0.50) | 0.779 (0.740, 0.815) |
| CovNet | 0.808<br>(0.775, 0.841) | 0.3 | 0.305 (0.247, 0.367) | 0.969 (0.951, 0.982) | 0.813 (0.718, 0.887) | 0.760 (0.727, 0.791) |
|  |  | 0.4 | 0.354 (0.294, 0.418) | 0.955 (0.934, 0.971) | 0.775 (0.686, 0.849) | 0.771 (0.737, 0.802) |
|  |  | 0.5 | 0.387 (0.325, 0.451) | 0.940 (0.917, 0.959) | 0.740 (0.655, 0.814) | 0.777 (0.744, 0.808) |
|  |  | 0.6 | 0.432 (0.369, 0.497) | 0.937 (0.913, 0.956) | 0.750 (0.670, 0.819) | 0.790 (0.756, 0.820) |
|  |  | 0.7 | 0.473 (0.409, 0.538) | 0.931 (0.907, 0.951) | 0.752 (0.675, 0.818) | 0.801 (0.768, 0.831) |
| Mixed-Effects<br>(Ours) | 0.930<br>(0.914, 0.947) | 0.3 | 0.675 (0.612, 0.733) | 0.935 (0.911, 0.954) | 0.820 (0.760, 0.871) | 0.867 (0.838, 0.894) |
|  |  | 0.4 | 0.741 (0.681, 0.795) | 0.904 (0.877, 0.927) | 0.773 (0.713, 0.825) | 0.888 (0.859, 0.913) |
|  |  | 0.5 | 0.778 (0.720, 0.828) | 0.882 (0.853, 0.908) | 0.744 (0.686, 0.797) | 0.90 (0.872, 0.924) |
|  |  | 0.6 | 0.827 (0.774, 0.873) | 0.859 (0.827, 0.887) | 0.720 (0.664, 0.772) | 0.919 (0.892, 0.941) |
|  |  | 0.7 | 0.885 (0.838, 0.922) | 0.828 (0.794, 0.859) | 0.694 (0.639, 0.744) | 0.942 (0.918, 0.961) |

Similar to the internal validation, Figure 7 shows reasonable calibration for the mixed-effects model, although some recalibration may improve performance. Again, the comparison models could give harmful predictions as they are poorly calibrated. The decision curve in Figure 8 shows that the model is of great clinical benefit compared to the treat all and treat-none approach.

**Figure 7:**
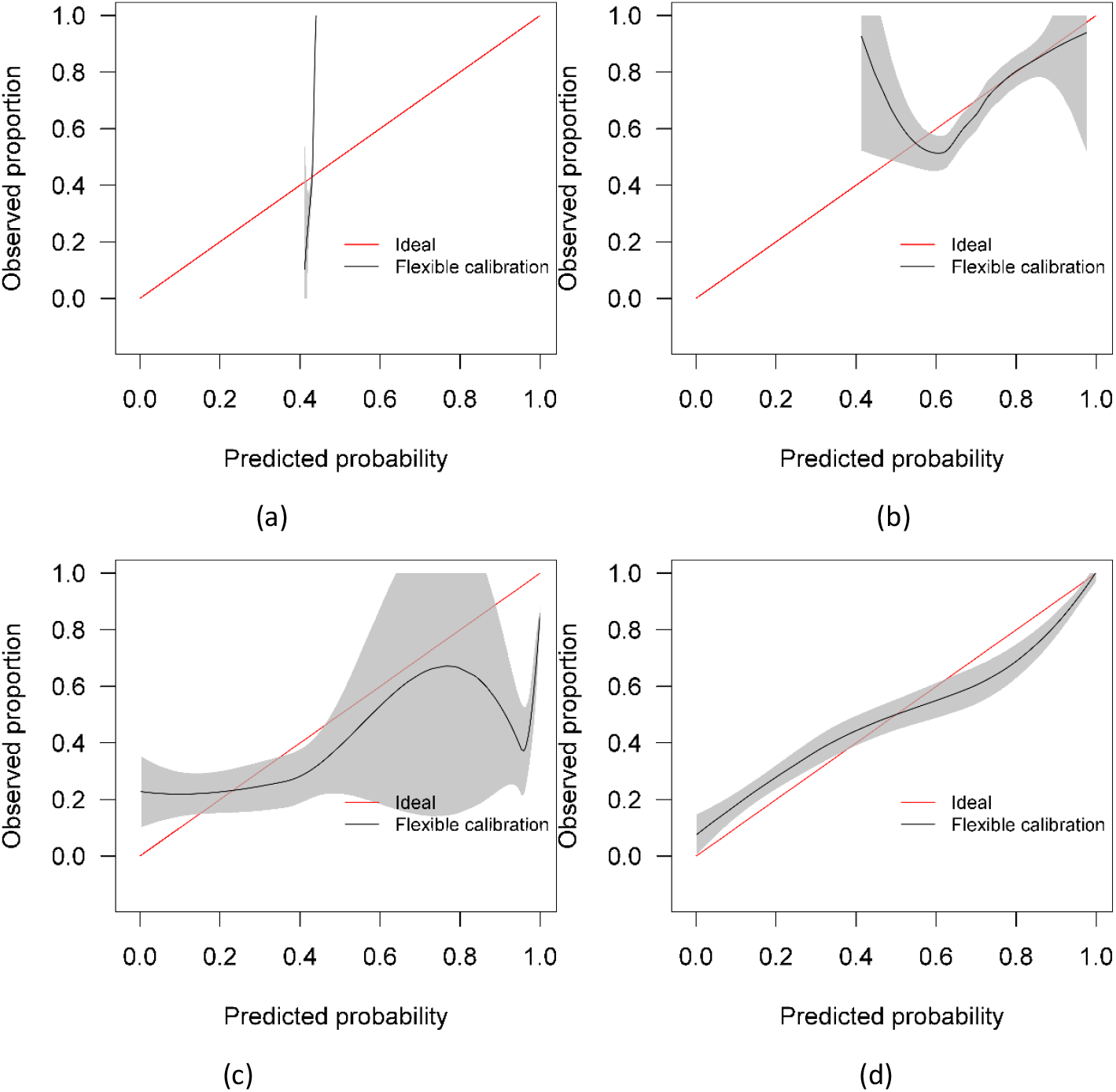
Calibration curves for (a) the Bai et al.^8^ model (b) the Covinet model^9^, (c) the Covnet model^7^, (d) the proposed mixed-effects model on the Zhang et al. external validation dataset.

**Figure 8:**
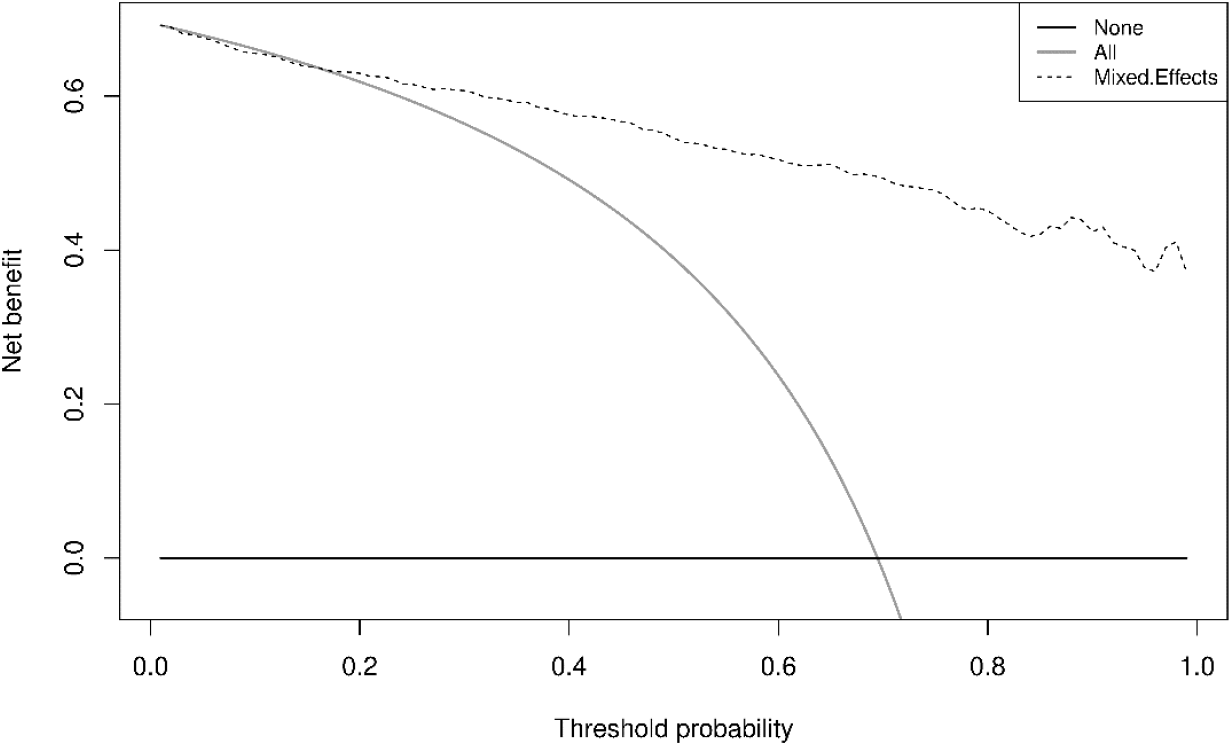
Decision curves for the proposed mixed-effects model on the Zhang et al. external validation dataset.

Although our proposed method and the Covnet model showed comparable performance on the internal validation set, the Covnet model could not generalise to the external geographical validation set, and calibration showed that the Covnet model would provide harmful risk estimates. This highlights the need for robust external validation in each intended setting. Nevertheless, the results show that the proposed method better generalises to external geographical datasets and provides less harmful predictions when compared to the four previously proposed methods based on the calibration curves.

### Saliency maps

It is vital to understand how the algorithm makes decisions and to check that it identifies the correct features within the image. Saliency maps can be used as a visual check to see what features the algorithm is learning. For example, the saliency maps in Figure 9 show that the model correctly identifies the diseased areas of the scans. We used 100 samples with a smoothing noise of 0.05 to create these saliency maps.

**Figure 9:**
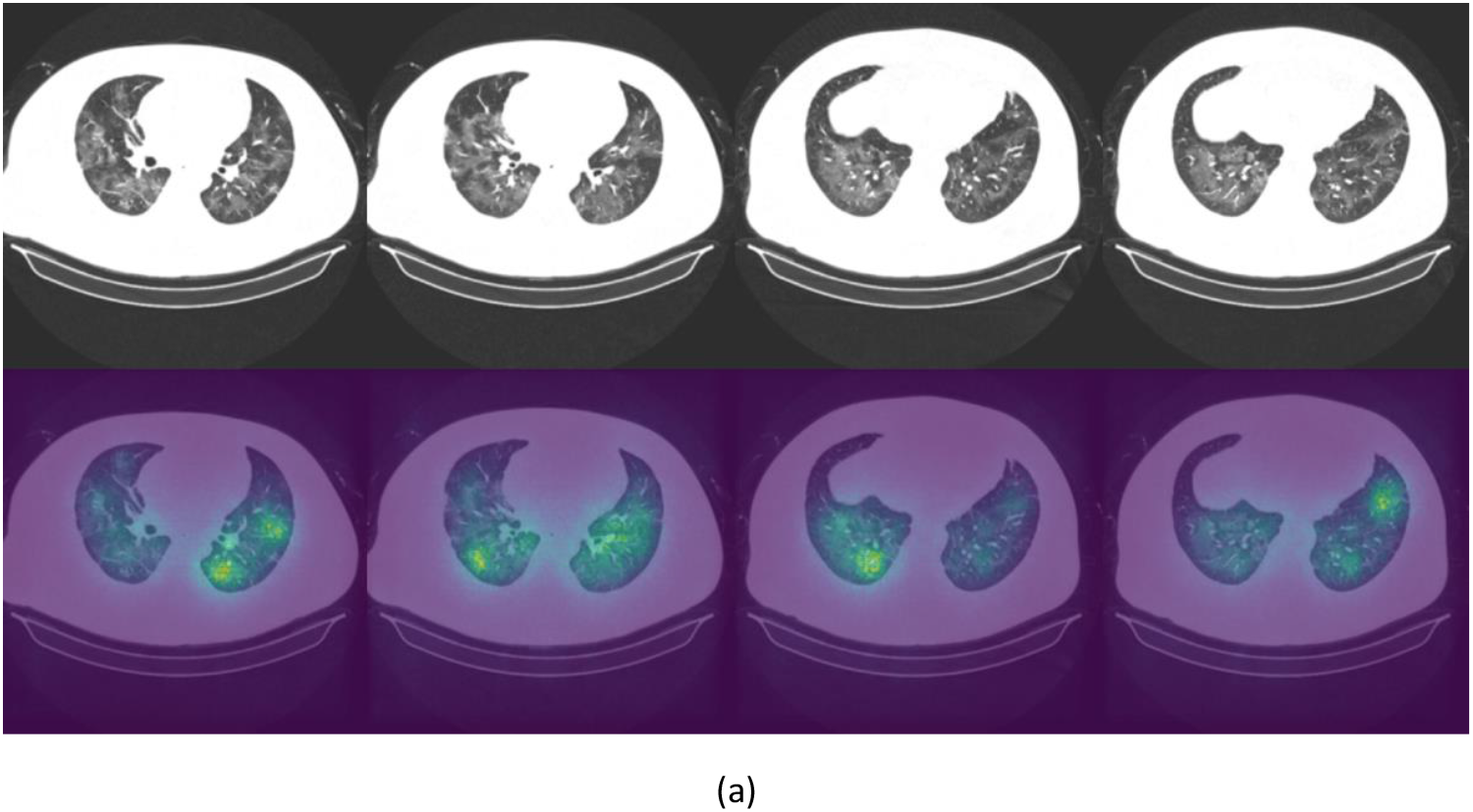
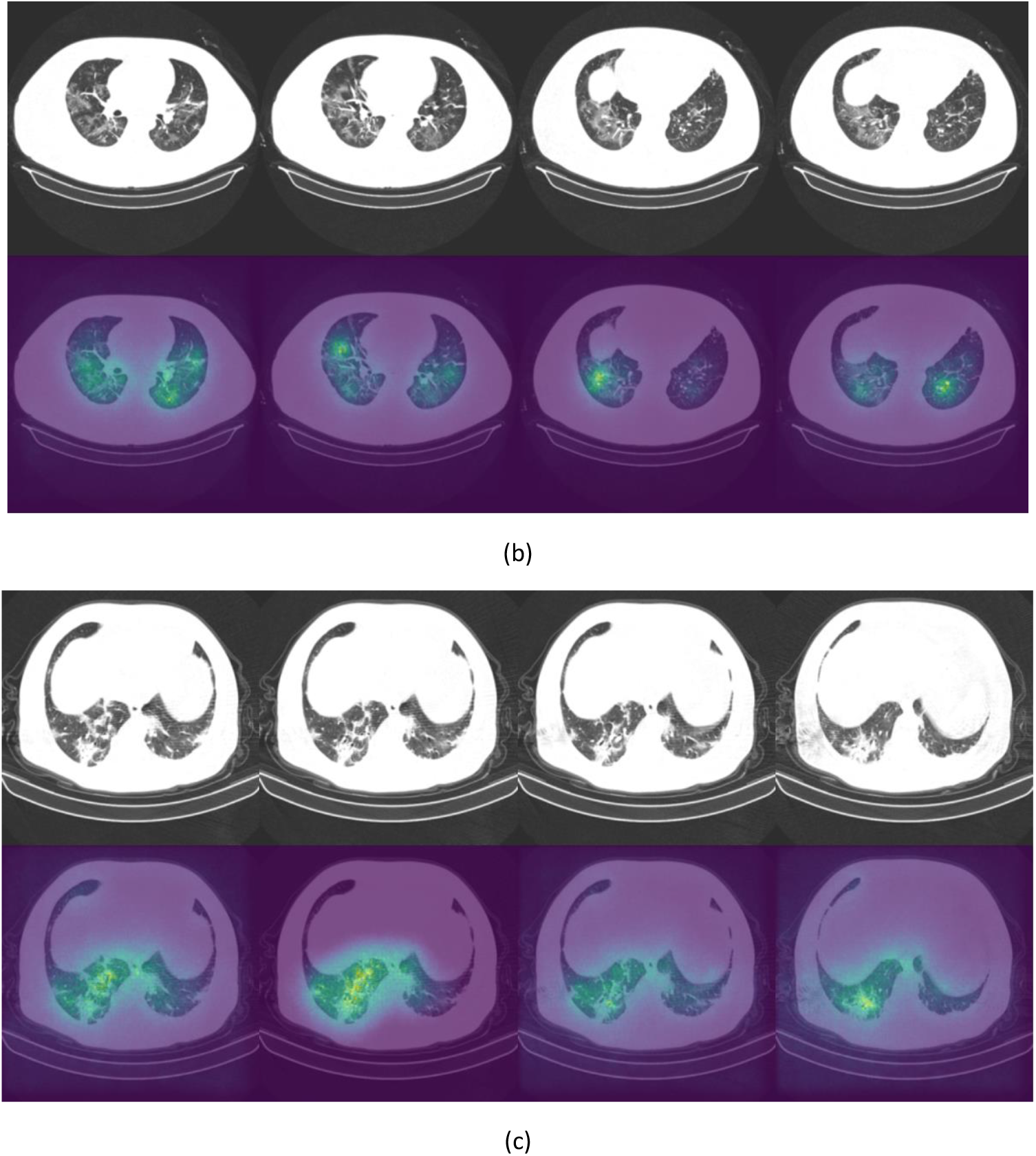
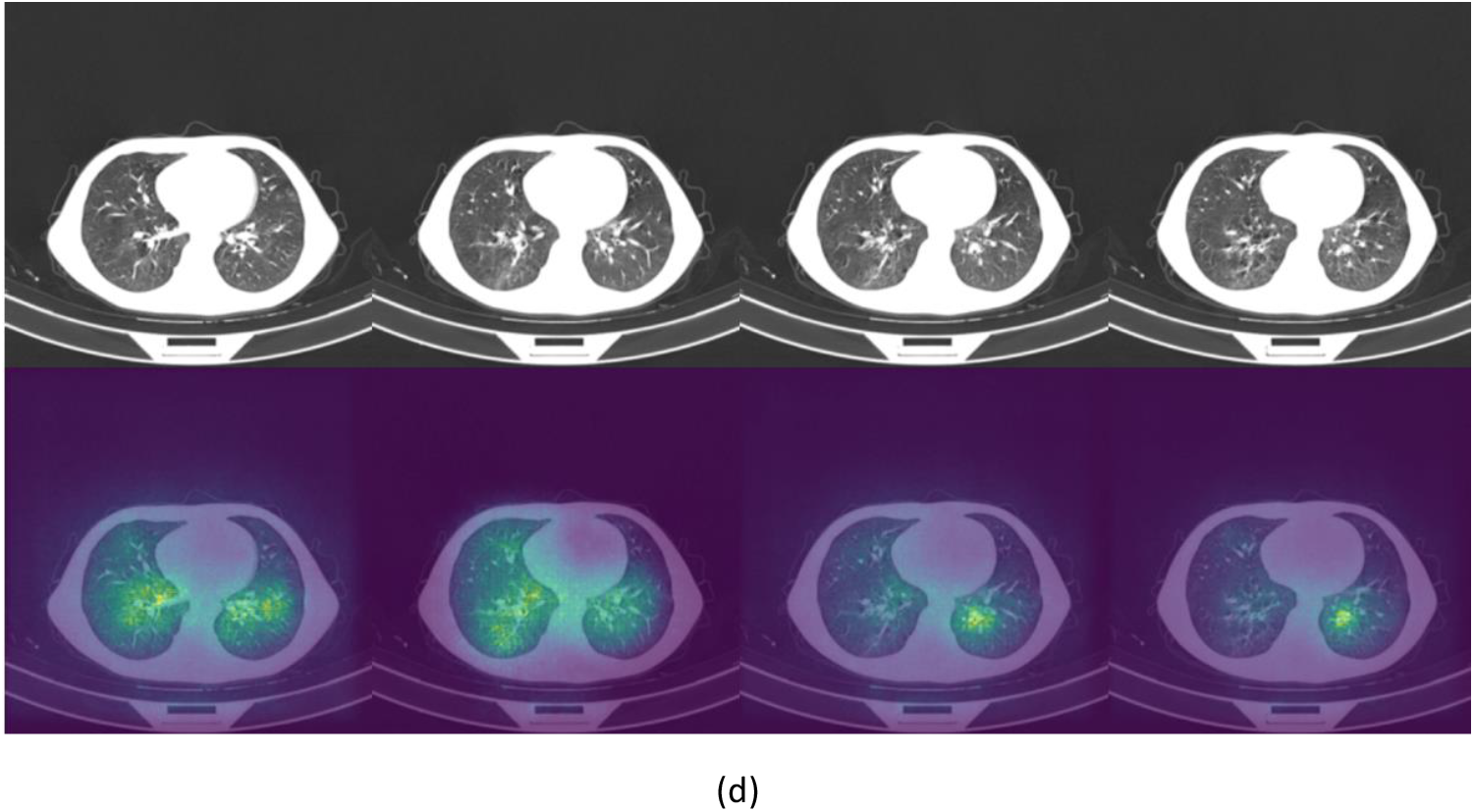
Example of original images and saliency maps showing highlighted regions on four patients (a, b, c, and d) in the Zhang et al.^48^ dataset. Four consecutive slices display how the diseased areas differ between slices. All images are taken from the external validation set.

### Sensitivity analysis

Mixed-effects models are capable of accounting for missing data. However, only one image had less than 20 slices; hence, we could not adequately assess if our model can indeed maintain good performance with missing data. Here, we rerun the analysis using the same dataset, using the same model and weights; however, we reduce the number of slices available as testing data inputs to simulate missing data. Blank images replace these slices. We uniformly sampled the slices choosing between 10 and 19 slices; this equates to between 5 and 50% missing data for the model. We ran inference at each level of missingness and briefly show the AUROC to determine at which point the predictive performance is significantly reduced.

The plot of AUROCs at different levels of missingness is shown in Figure 10, along with 95% confidence intervals. We can see that at 20% missingness, there is a statistically significant decrease in predictive performance. Although, even at 50% missingness, the model still performs relatively well, with an AUROC of 0.890 (95% CI: 0.868, 0.912). It should be noted that this does not mean that there is no reduction in performance at 5-15% missingness, only that the reduction was not statistically significant at the 95% confidence level.

**Figure 10:**
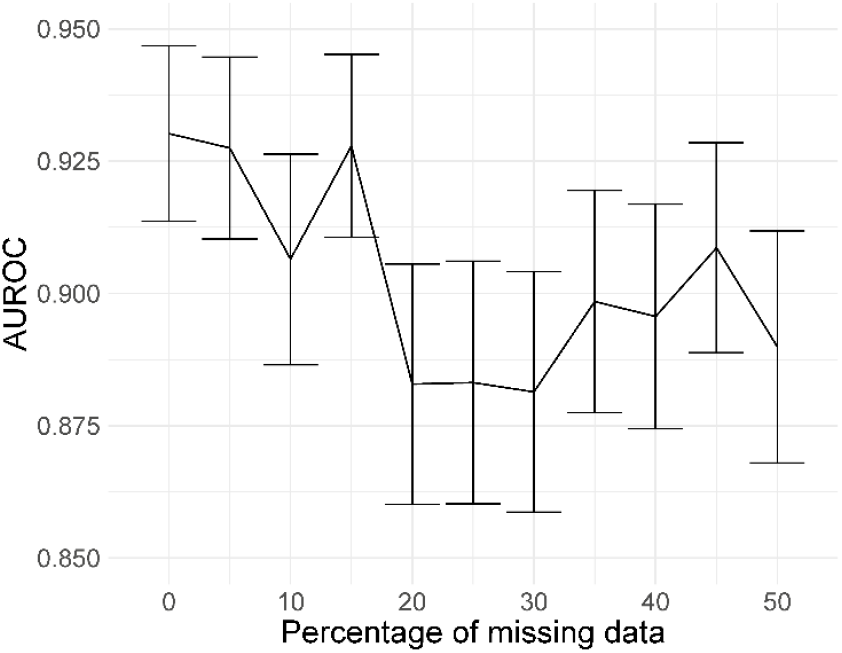
AUROC values at different levels of missingness. At 20% missingness, the loss in performance becomes statistically significant; however, even with 50% missing images, the model still has a reasonably high AUROC.

Deep learning models can be susceptible to adversarial attacks^49^, where minor artefacts or noise on an image can cause the image to be misclassified, even when the image does not look significantly different to a human observer. Here, we perform a brief sensitivity analysis by adding a small Gaussian noise to the image. We tested the model performance on the external dataset, with each image having a random Gaussian noise added. Experiments were conducted with standard deviations of 0 up to 0.005 in increments of 0.001 added to the normalised image. We did not add Gaussian noise in the data augmentation so that the model is not explicitly trained to deal with this kind of attack.

When using a variance of 0, the images are unchanged, and the results are the same as the standard results above. We present results on the Zhang et al.^48^ dataset. Example images for each level of variance are shown in Figure 11, and a graph showing the reduction in AUROC is shown in Figure 12.

**Figure 11:**
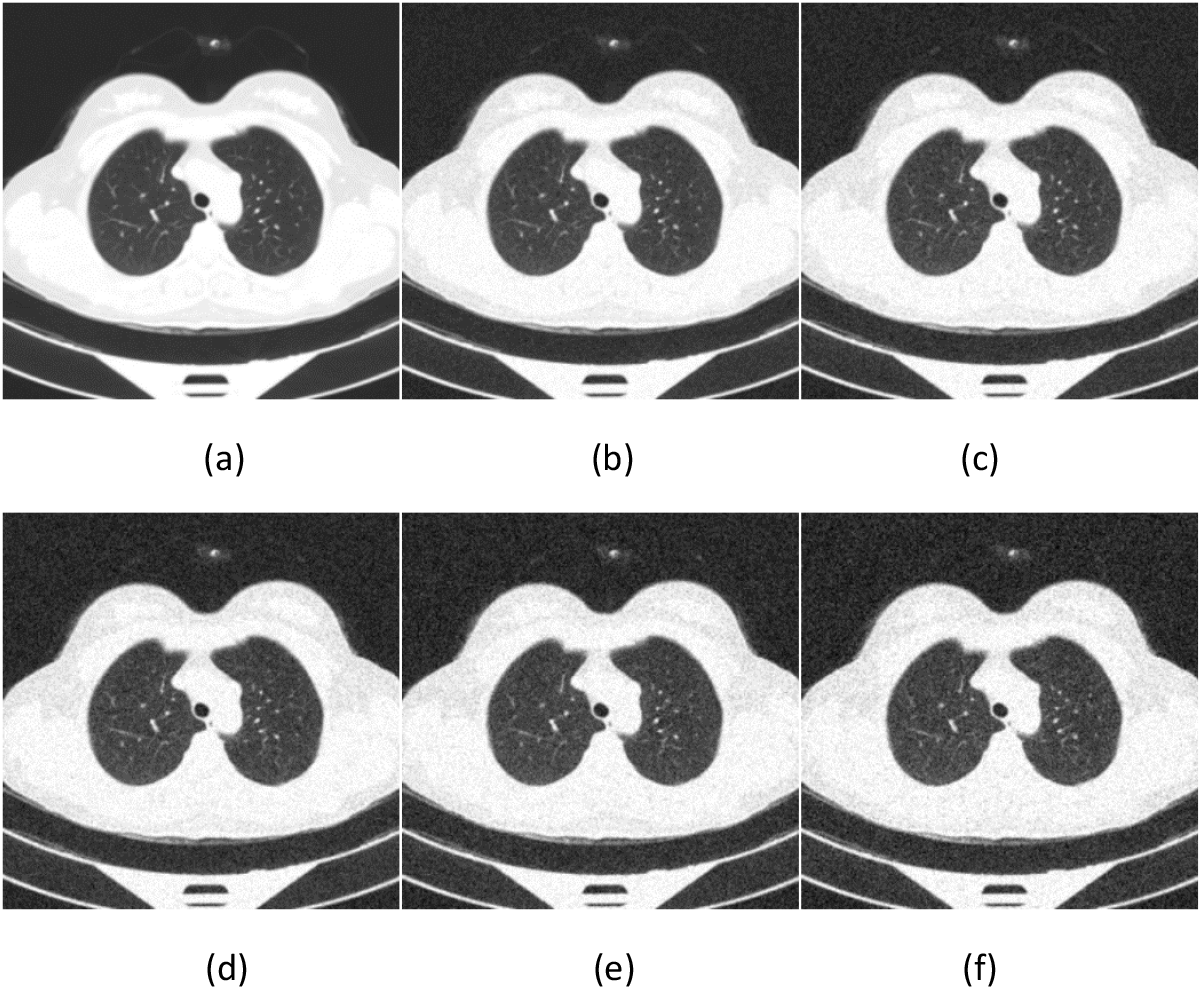
Example images showing the effect of increasing the amount of noise in the image input. Noise is increased by increasing the standard deviation (s.d.) of the Gaussian noise. (a) s.d.=0; (b) s.d.=0.001; (c) s.d.=0.002; (d) s.d.=0.003; (e) s.d.=0.004; (f) s.d.=0.005.

**Figure 12:**
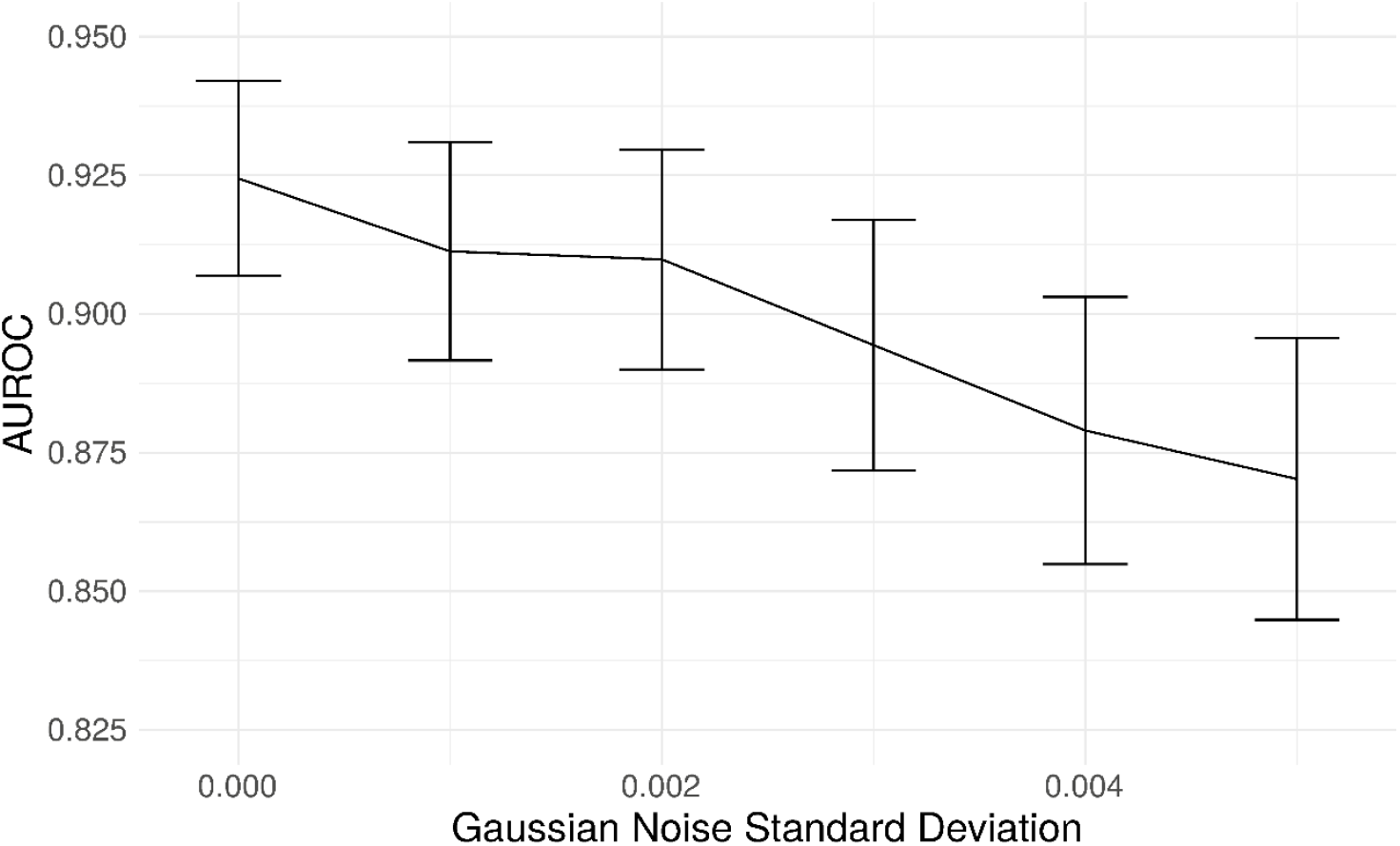
Graph showing the drop in AUROC as the amount of noise in the image input increases. The AUROC falls steadily with increased noise in the image.

## Discussion

Artificial intelligence is set to revolutionise healthcare, allowing large amounts of data to be processed and analysed automatically, reducing pressure on stretched healthcare services. These tools can aid clinicians in monitoring and managing both common conditions and outbreaks of novel diseases. However, these tools must be assessed adequately, and best practice guidelines for reporting and development must be followed closely to increase reproducibility and reduce bias. We have developed a deep learning model to classify CT scans as healthy or COVID-19 using a novel mixed-effects model. Following best practice guidelines, we have externally validated the model. In addition, we robustly externally geographically validated the developed model in several performance areas, which are not routinely reported. For example, discriminative performance measures show that the model can discriminate between healthy and COVID-19 CT scans well, calibration shows that the model is not clinically harmful. Finally, the clinical usefulness measures show that the model may be useful in a clinical setting. From the results presented here, it would seem that our deep learning model outperforms the RT-PCR tests as shown in the review by Watson et al. ^3^; however, those results are conservative estimates and were conducted under real-world clinical settings. A prospective study is required to determine if this is the case.

Compared to previously proposed models, our model showed similar discriminative performance to one existing method; however, our method generalised better to an external geographical validation set and showed improved calibration performance. Interestingly, in both internal and external validation, the sensitivity and NPV are similar in all models. However, the specificity and PPV are statistically significantly improved for the mixed-effects model in the external validations dataset. The performance of the proposed model in the external validation set is similar to that reported by PCR testing^3^. However, a direct comparison should not be made as PCR testing on this exact dataset is unavailable.

There are several limitations of the study that should be highlighted and improved in future work. Firstly, we have only performed external geographical validation in a single dataset. Further external validation, both geographical and temporal, is needed on many datasets to determine if the model is correct in each intended setting. Although we performed a brief sensitivity analysis here, more extensive work on adversarial attacks is needed. Future studies could consider following the method proposed by Goodfellow et al.^49^ to improve robustness against adversarial examples. Patient demographic data were not available for this study, but future studies could incorporate this data into the model to improve results. Finally, rules of thumb for assessing sample size calculations in the validation set can lead to imprecise results^50^. Simulating data is a better alternative; however, it is difficult to anticipate the distribution of the model’s linear predictor. Therefore, we were required to revert to the rule of thumb using a minimum of 200 samples in each group^28^.

Initial experiments used the Zhang et al.^48^ dataset for training; this showed promising results on the internal validation set; however, external validation showed random results. In addition, saliency maps showed that the model was not using the features of COVID-19 to make the diagnosis and was instead using the area around the image. We concluded that the images for each class were slightly different, perhaps due to different imaging protocols, and the algorithm was learning the image format rather than the disease. We then used the MosMed dataset for training and the Zhang et al.^48^ dataset for external validation. This highlights the need for good quality training data and external validation and visualisation.

Future studies should validate models and follow reporting guidelines such as TRIPOD^17^ or the upcoming QUADAD-AI^51^ and TRIPOD-AI^52^ to bring about clinically useful and deployable models. Further research could look deeper into the areas of images identified by the algorithm as shown on the saliency maps; this could potentially identify new features of COVID-19 which have gone unnoticed. Before any model can be fully deployed, clinical trials are needed to study the full impact of using such algorithms to diagnose COVID-19 and the exact situations in which such a model may be used. In-clinic prospective studies comparing the performance deep learning models with RT-PCR and lateral flow tests should be carried out to determine how deep learning compares; this will show whether deep learning could be used as an automated alternative to RT-PCR testing.

This study indicates that deep learning could be suitable for screening and monitoring of COVID-19 in a clinical setting; however, validation in the intended setting is vital, and models should not be adopted without this. It has been observed that the quality of reporting of deep learning prediction models is usually very poor; however, with a bit of extra work and by following best practice guidelines, this problem can be overcome. This study highlights the importance of robust analysis and reporting of models with external validation.

## Data Availability

All data used is publicly available from the following links: https://mosmed.ai/datasets/covid19_1110/ http://ncov-ai.big.ac.cn/download?lang=en

